# High Prevalence of Unrecognized Actionable Cardiac Arrhythmias in Patients with Moderate to Severe Chronic Obstructive Pulmonary Disease

**DOI:** 10.1101/2024.10.11.24315304

**Authors:** Kristie M. Coleman, RN. Elliot Wolf, Dimitrios Varrias, Jacob Schwartz, Brenda Garcia, Victoria Roselli, Betty Lam, Jonas Leavitt, Nikhil Sharma, Gregory Dumchin, Erica Altschul, Margarita Oks, Bushra Mina, Stavros E. Mountantonakis

**Affiliations:** Lenox Hill Hospital; Lenox Hill Hospital, Northwell Cardiovascular Institute; Hofstra Northwell School of Medicine; Lenox Hill Hospital, Northwell Health

**Keywords:** Chronic obstructive pulmonary disease, Arrhythmias, Long-term continuous monitoring

## Abstract

**Background:** Patients with chronic obstructive pulmonary disease (COPD) are at high risk for developing arrhythmias due to hypoxemia, right heart failure, and the use of beta-agonist inhalers. Symptoms related to arrhythmias can often be masked or confounded by symptoms related to COPD exacerbation and remain undiagnosed. With this study, we identify the incidence of actionable arrhythmias in patients with no prior cardiology follow-up and moderate-severe COPD with continuous monitoring.

**Methods:** An automatic referral for electrophysiology (EP) consult was generated in patients with moderate-severe COPD if they endorsed one of the following: palpitations, dizziness, abnormal ECG, or near syncope. Eligible patients underwent ILR implantation after evaluation with an EP specialist and were followed via remote monitoring for 12 months. A control group of patients without COPD matched for age, sex, and implant indication were randomly selected in a 3:1 ratio. Actionable arrhythmias, defined as arrhythmias that correlated with symptoms triggered by the patient, necessitating EP intervention, were recorded for both groups.

**Results:** In this prospective cohort study, 21 patients with COPD were enrolled and compared to 63 controls. COPD patients experienced a significantly higher rate of actionable arrhythmias compared to the controls (48% vs 11%, p<0.001). EP interventions in response to actionable arrhythmias included eight patients initiated on anticoagulation, three catheter ablations, one implantable cardiac defibrillator, and one permanent pacemaker implanted. In multivariate analysis, COPD was an independent predictor of actionable arrhythmias (aOR 4.3, 95% CI 1.2-15.2, p=0.02) when adjusting for chronic kidney disease and all-cause readmissions.

**Conclusion:** Continuous monitoring was highly effective in diagnosing significant arrhythmic events in patients with moderate-severe COPD. Awareness should be raised about the high arrhythmic risk in this population and the role of continuous monitoring should be evaluated in larger studies.

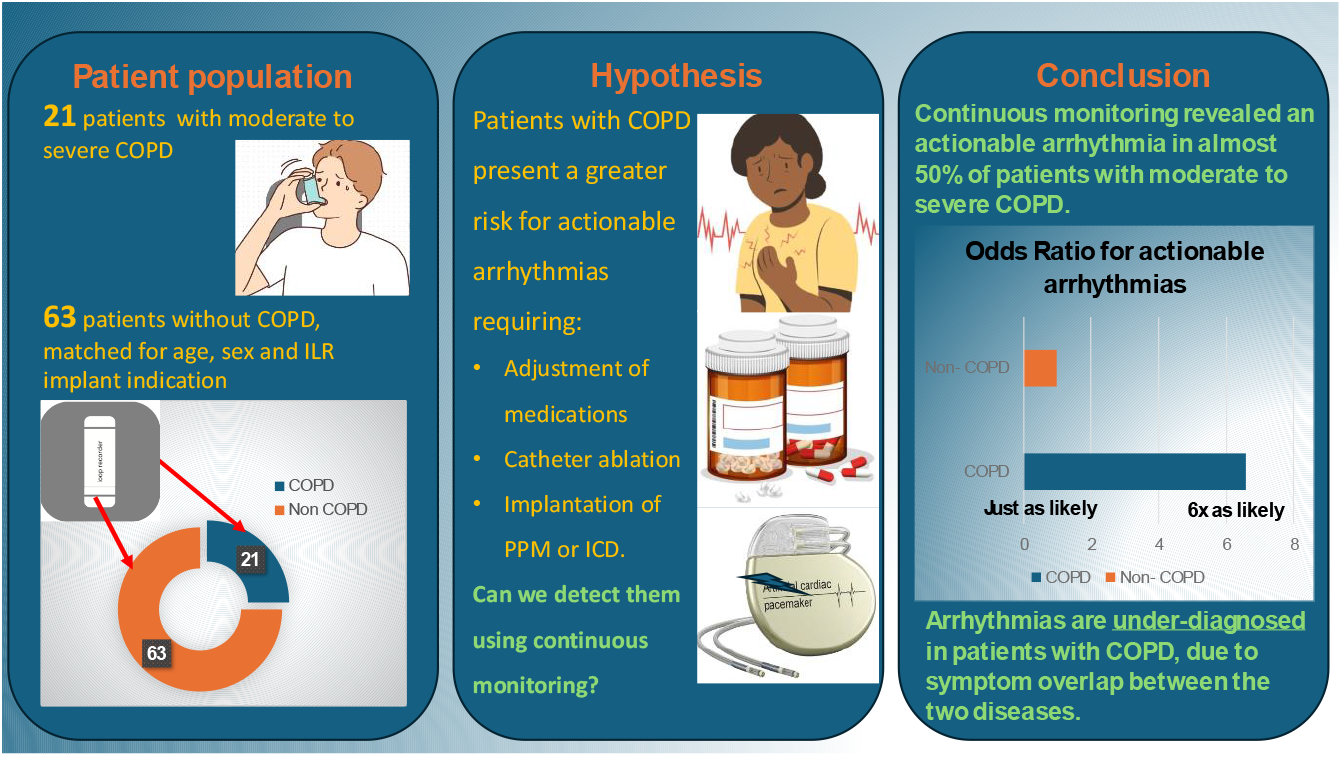

**WHAT IS KNOWN?:**

- Patients with COPD are at greater risk for developing cardiac arrhythmias, which may propagate COPD exacerbations.

**WHAT THE STUDY ADDS:**

- Quantification of incidence of actionable arrhythmias in patients with moderate to severe COPD using continuous monitoring for the 1^st^ time.
- Multivariate analysis which determines whether this phenomenon is due to demographic confounders, comorbidities, treatment modalities or an independent association.
- Detailed presentation of the type or arrhythmias, COPD exacerbations and healthcare utilization, emphasizing the need for arrhytmia surveillance in this vulnerable patient population.

## INTRODUCTION

Atrial arrhythmias have a significantly higher incidence in patients with chronic obstructive pulmonary disorder (COPD) compared to the general population [1]. The incidence of atrial fibrillation (AF), in particular, is 1.8 times more common in COPD and tends to be refractory to medical and ablative therapies [2]. In addition, cardiovascular events are a leading cause of death in COPD patients, with new studies suggesting arrhythmias and sudden cardiac death as substantial contributors [3]. COPD patients are at a higher risk for ventricular tachycardia (VT) and mortality [4] and the frequency of inappropriate implantable cardioverter-defibrillator (ICD) shocks is higher in COPD patients compared to non-COPD patients [5-13].

Given the complex interplay of factors that may predispose COPD patients to arrhythmogenesis, understanding the relationship between COPD and arrhythmia could help physicians better identify those who could benefit from an electrophysiology intervention [14]. The true incidence of silent arrhythmias has not been thoroughly investigated; previous retrospective studies have sought to do so using 24-hour Holter monitoring [15]. However, sustained arrhythmias (ventricular or atrial fibrillation) were rarely captured within the short monitoring period of 24 hours, limiting arrhythmia risk assessment to only non-sustained VT [16, 17]

Our study aimed to identify the incidence of actionable arrhythmias in patients with no prior cardiology follow-up and moderate-severe COPD with continuous monitoring compared to a control group matched for age, sex, and implant indication. We also sought to examine the association between COPD and actionable arrhythmias.

## METHODS

### Study Design and Population

In this prospective cohort study, patients were recruited for enrollment through a collaborative outreach effort between the pulmonary and electrophysiology (EP) teams. An automatic referral for EP consult was generated in patients aged 18-75 years with moderate-severe COPD as defined as Global Initiative for Chronic Obstructive Lung Disease (GOLD) Stage 2-4 if they answered yes to one of the following: palpitations, dizziness, abnormal ECG, near syncope during a routine pulmonary clinic visit. Patients with a life expectancy of less than one year, a history of documented arrhythmic syndrome (including atrial fibrillation, supraventricular tachycardia, frequent premature ventricular complexes, ventricular tachycardia or ventricular fibrillation), a history of sudden cardiac death, and an existing pacemaker or implantable cardiac device were excluded. Upon consultation with an EP specialist, the implantation of a loop recorder (ILR) was discussed with the patient, and if agreeable, the patient underwent ILR implantation. Patients were monitored via remote home monitoring for 12 months post ILR implantation. Patients were also asked to log potential symptoms of clinical arrhythmias. This study received approval from the Northwell Human Research Protection Program, consent was obtained from study participants.

### Comparator Group

A retrospective control cohort matched for age, sex, and implant indication was randomly selected in a 3:1 ratio from a computer-generated list of patients who received ILRs at Lenox Hill Hospital and had at least 12 months of data post-implant.

### Data Sources

Patients’ baseline characteristics, including demographics, comorbidities, medications, ECG measurements, and pulmonary function test (PFT) results for the COPD group, were recorded. Patients were then followed for the study duration via remote monitoring. Arrhythmia data were evaluated via monthly review of transmissions for events meeting the protocol-defined arrhythmia threshold.

Patient’s electronic medical records were reviewed to evaluate the occurrence of EP interventions and general healthcare utilization. Concurrently, the patients were monitored for any incidence of COPD exacerbation.

### Clinical Events and Outcomes

#### Actionable Arrhythmias

Actionable arrhythmias were defined as arrhythmias that correlated with symptoms triggered by the patient, necessitating adjustment of medications (including initiation of anticoagulation), catheter ablation and implantation of implantable cardiac defibrillator (ICD) or permanent pacemaker (PPM). Atrial arrhythmia was defined by a single AF episode lasting longer than one hour or a cumulative daily AF burden of 5% or greater. Ventricular arrhythmia was defined as a sustained arrhythmia lasting more than thirty seconds or a cumulative daily PVC burden of 5 % or greater. Conduction abnormalities noted via remote monitoring were recorded.

#### COPD Exacerbation

Acute COPD exacerbation was defined as an acute worsening of respiratory symptoms that resulted in additional therapy. Exacerbations were classified as 1) mild if they are treated with short-acting bronchodilators only; 2) moderate if they are treated with short-acting bronchodilators plus antibiotics and/or oral corticosteroids; or 3) severe if the patient visits the emergency room or requires hospitalization because of an exacerbation.

#### Healthcare Resource Utilization

The frequency and proportion of cardiovascular and all-cause admissions within the 12-month study period between the two groups was evaluated, mortality was also recorded.

### Statistical Analyses

Normally distributed data are presented as mean ± SD and non-normal data as median (interquartile range [IQR]). Categorical data are presented as frequency (percentage of the total). Continuous variables were compared using the parametric Student *t* test or nonparametric Mann-Whitney U test. Categorical variables were compared using the χ^2^ test or Fisher’s exact test when greater than 20% of the cells had expected frequencies less than 5. To evaluate the association of COPD with actionable arrhythmias, a stepwise logistic regression analysis was performed to examine variables associated with actionable arrhythmias with the prespecified level of significance for removal (*P* >.20) and for entry (*P* < .20). Adjusted odds ratios (AORs) and 95% CIs were obtained.

Study data were collected and managed using REDCap electronic data capture tools hosted at Northwell Health. The data analysis for this paper was generated using STATA 17.0 (Statacorp. 2021. Stata Statistical Software: Release 17. College Station, TX: StataCorp LLC).

## RESULTS

### Patient Characteristics

In the prospective COPD cohort, 21 patients were enrolled and 63 patients without a prior diagnosis of COPD, matched for age, sex and implant indication were included in the control group. Clinical and demographic characteristics of the patient population are listed in Table 1 for both the COPD and control groups.

**TABLE 1:** Baseline characteristics.

|  |  | Control (n = 63) | COPD (n = 21) | p-value |
| --- | --- | --- | --- | --- |
| **Sex = male** |  | 23 (37%) | 10 (48%) | 0.37 |
| **Age (years), mean (± SD)** |  | 65.48 (± 14.61) | 69.38 (± 9.73) | 0.26 |
| **Race** | White | 39 (62%) | 18 (86%) | 0.29 |
|  | Black/African American | 15 (24%) | 3 (14%) |  |
|  | Asian | 3 (5%) | 0 (0%) |  |
|  | Other/Multiracial | 4 (6%) | 0 (0%) |  |
|  | Unknown | 2 (3%) | 0 (0%) |  |
| **BMI, mean (± SD)** |  | 28.35 (± 7.34) | 27.75 (± 6.02) | 0.74 |
| **Smoking Status** | Current smoker | 1 (2%) | 6 (29%) | <0.001 |
|  | Former smoker | 14 (22%) | 10 (48%) |  |
|  | Never smoker | 48 (76%) | 5 (24%) |  |
| **Medical History** | Heart surgery | 3 (5%) | 0 (0%) | 0.31 |
|  | PCI/angioplasty | 8 (13%) | 3 (14%) | 0.85 |
|  | Diabetes mellitus | 7 (11%) | 4 (19%) | 0.35 |
|  | Hypertension | 39 (62%) | 15 (71%) | 0.43 |
|  | Hyperlipidemia | 31 (49%) | 12 (57%) | 0.53 |
|  | Heart failure | 3 (5%) | 0 (0%) | 0.31 |
|  | Coronary artery disease | 11 (17%) | 5 (24%) | 0.52 |
|  | Myocardial infarction | 4 (6%) | 2 (10%) | 0.62 |
|  | Chronic kidney disease | 3 (5%) | 4 (19%) | 0.04 |
|  | Stroke/TIA | 7 (11%) | 5 (24%) | 0.15 |
|  | Non-long cancer | 10 (16%) | 4 (19%) | 0.74 |
|  | Asthma | 2 (3%) | 5 (24%) | 0.003 |
|  | Obstructive sleep apnea | 4 (6%) | 4 (19%) | 0.09 |
| **Medication Use** | Beta blocker | 29 (46%) | 9 (43%) | 0.80 |
|  | Calcium channel blocker | 3 (5%) | 3 (14%) | 0.14 |
|  | Anticoagulant | 5 (8%) | 3 (14%) | 0.39 |
|  | Antiplatelet | 30 (48%) | 8 (38%) | 0.45 |
|  | Antiarrhythmic drug | 3 (5%) | 0 (0%) | 0.31 |
|  | Diuretic | 14 (22%) | 7 (33%) | 0.31 |
|  | Statin | 35 (56%) | 11 (52%) | 0.80 |
|  | ACE Inhibitor | 9 (14%) | 3 (14%) | .99 |
|  | Angiotensin receptor blocker | 18 (29%) | 5 (24%) | 0.67 |
| **ILR Indication** | Near Syncope | 10 (15.9%) | 3 (14.3%) | 0.86 |
|  | Abnormal ECG | 7 (11.1 %) | 2 (9.5%) | 0.84 |
|  | Palpitations | 19 (30.2%) | 8(38.1%) | 0.50 |
|  | Dizziness | 27 (42.9%) | 8 (38.1%) | 0.70 |
| **EKG Parameters** | HR (bpm), mean (± SD) | 72.37 (± 13.73) | 79.29 (± 16.09) | 0.06 |
| | QTc (msec), mean (±SD) | 420.91 (± 25.07) | 427.71 (± 17.23) | 0.26 |
| | LVEF, mean (± SD) | 0.59 (± 0.09) | 0.61 (± 0.06) | 0.33 |

The two groups showed no difference in baseline characteristics or comorbidities, except chronic kidney disease (19% vs 5%, p=0.04) and asthma (24% vs 3%, p=0.003) were significantly higher in the COPD group. Expectedly, a significantly larger proportion were current or former smokers in the COPD group (76% vs 24%, p < 0.001).

### Clinical Events and Outcomes

#### Actionable Arrhythmias

The incidence of actionable arrhythmias in COPD patients was significantly higher compared to the controls (48% vs 11%, p<0.001). The incidence of atrial arrhythmias was significantly higher in the COPD cohort, with 43% of COPD patients experiencing atrial arrhythmias compared to 13% in the control group (p = 0.003). There was no correlation between patient reported symptoms and documented clinical arrhythmias. Initiation of anticoagulation in response to incident AF was significantly higher in the COPD group (38.0% vs 3.0%, p<0.001). Three out of 21 patients (14%) from the COPD group underwent catheter ablation as a result of arrhythmia monitoring, as opposed to 5 out of 63 (8.0%) patients in the control group.

Ventricular arrhythmias were also more common in the COPD cohort, occurring in 4(19%) of COPD patients compared to 2(3%) in the control group (p = 0.014). In the COPD cohort, 2 patients experienced sustained ventricular tachycardia, with one patient undergoing ILR extraction and implantable cardiac defibrillator implantation in response. Ventricular ectopy >5% was noted in the remaining patients, with an average PVC burden of 13.9% in the COPD patients and 6.1% in the control. There was no statistically significant difference in time to incident ventricular or atrial arrhythmia throughout the monitoring period.

Conduction system disease was documented in 2 patients in the control group compared to 1 in the COPD group (3.2% vs 4.8%, p=0.73). Permanent pacemakers were implanted for tachy-brady syndrome and complete heart block in the control patients, respectively, and for sinus node dysfunction in the COPD patients.

A multivariate logistic regression analysis, adjusting for age, sex, BMI, CKD, smoking status, diabetes, hypertension, and number of readmissions, revealed that COPD (aOR 6.6, 95% CI 1.9-23, p=0.03) was independently associated with an increased incidence of actionable arrhythmias. None of the other variables reached statistical significance levels. (Figure 1.)

**Figure 1.**
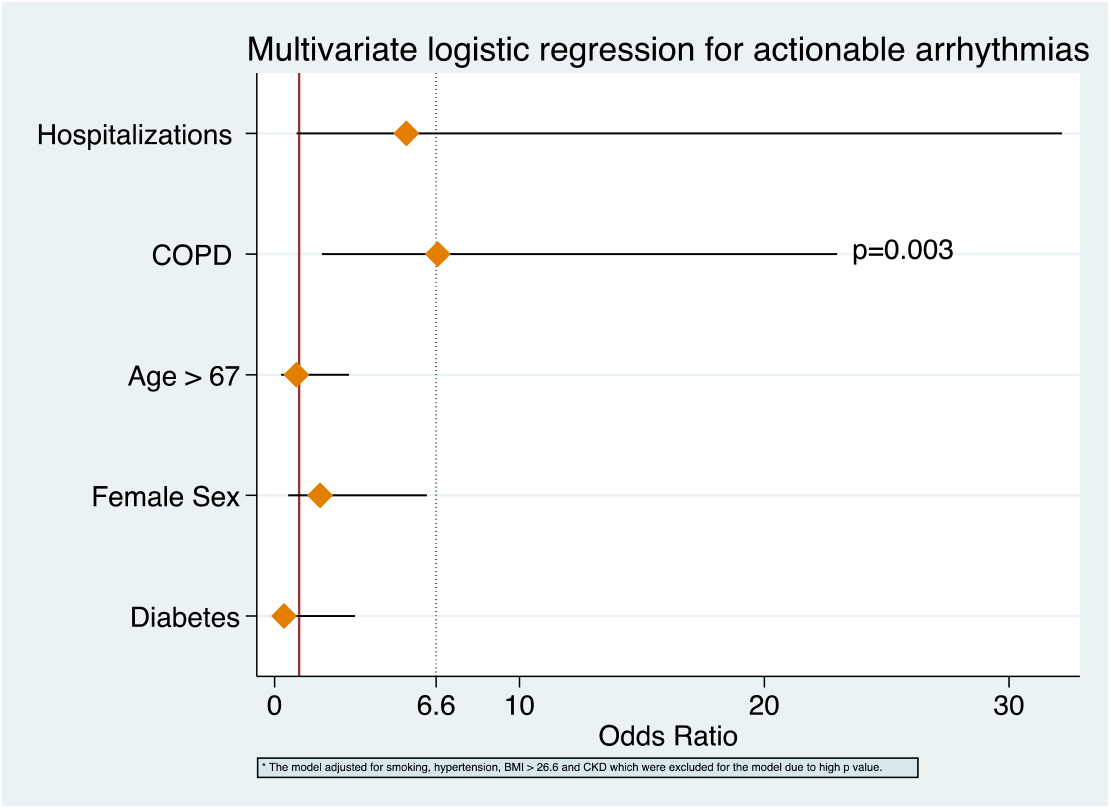
Logistic regression model predicting actionable arrhythmias using comorbidities. COPD is independently associated with increased risk.

#### COPD Exacerbations

Throughout the monitoring period, 7 COPD patients experienced exacerbations, categorized as moderate (2), severe (4), and mild (1). The use of short (SABA) and long-acting (LABA) beta-agonists was measured and found not to be associated with the incidence of actionable arrhythmias. In fact, 4/14 (28.6%) of patients with LABA had an actionable arrhythmia, compared to 5/8(62.5%) who did not have an event on LABA.

#### Healthcare Resource Utilization

Patients in the COPD group experienced a higher rate of cardiac-related admissions compared to the control (19.1% vs. 6.3%, p=0.08), as well as all-cause readmissions (61.9% vs. 17.4%, p<0.001) within the 12-month study period. In the COPD group, three patients died compared to none in the control group (p = 0.002). The etiology of death in all three patients was attributed to sequelae from severe COPD exacerbations resulting in respiratory failure.

## DISCUSSION

With this prospective cohort study utilizing long-term continuous monitoring, we showed that 48% of patients with COPD had an actionable arrhythmia within the 12-month study period. Eight patients were initiated on oral anticoagulation for stroke prophylaxis, three patients underwent catheter ablation for atrial fibrillation, and two patients underwent implantation of cardiac implantable electronic devices. Most importantly, we showed COPD is associated with six times the risk of experiencing an actionable arrhythmia. Notably, the patients enrolled in our prospective cohort were recruited through an automatic referral mechanism created by the EP team to identify COPD patients whose overlapping symptomatology potentially masked undiagnosed clinical arrhythmias. In routine clinical practice, this population is not typically referred to cardiology, nor EP, emphasizing the need for increased awareness and surveillance.

Although there are many potential mechanisms behind the findings presented above, autonomic dysfunction seems to play a central role in the pathogenesis of both COPD and arrhythmogenesis. Patients with COPD are known to have functional alterations of cardiac autonomic modulation, accounting for reported elevations in heart rate, reduced baroreflex sensitivity, and reduced heart rate variability. Sympathetic overactivity and vagal impairment are widely recognized as prognostic indications for the development of AF [18]. The prevalence of arrhythmias and particularly AF in patients with COPD is vastly underestimated [19]. In our cohort, we found patient reported symptoms did not correlate with clinical arrhythmias, suggesting patients may misinterpret their arrhythmia related symptoms as COPD symptoms.Common clinical factors that exist in patients with COPD, especially during an exacerbation, including hypoxemia, metabolic disturbances, and use of beta-agonists, are well-established precipitating factors for arrhythmogenesis [19]. However, in our study, beta-agonist use was not independently associated with arrhythmic events. Furthermore, commonly coexisting comorbid conditions, including obstructive sleep apnea (OSA), pulmonary hypertension, obesity, and smoking, further increase the risk of arrhythmias and complicate the management of these patients [19].

COPD has been identified as an independent predictor of AF progression from paroxysmal to persistent and recurrence post-catheter ablation [20-22]). Mendez-Bailon et al. analyzed the Spanish National Hospital Discharge Database between 2004-2013 and found patients admitted for AF with concomitant COPD had lengthier stays, higher rates of in-hospital mortality, and were less likely to receive EP interventions [23]. Durheim et al. enrolled 1605 AF patients in their prospective registry with COPD and found a higher burden of symptoms, reduced quality of life, higher risk of all-cause mortality, and cardiovascular hospitalizations[24].

While the association between COPD and AF is widely accepted, there are no prior studies that have utilized long-term continuous monitoring in patients with COPD for surveillance of clinical arrhythmias. Over a third of the patients enrolled in our small cohort had AF detected by the ILR, necessitating initiation of anticoagulation. Multiple studies have reported the increased thromboembolic risk among patients with COPD, which Shen et al. reported was further increased (HR 1.63) by concomitant AF [25]. Detection of atrial fibrillation in patients with COPD is imperative to initiating anticoagulation for stroke prophylaxis and may support long-term arrhythmia surveillance. Furthermore, three patients underwent successful catheter ablation for symptomatic atrial fibrillation. A sub-analysis of the global multicenter GLORIA-AF registry reported COPD patients were 12% less likely to receive catheter ablation or cardioversion [26]. While concern over the efficacy of EP interventions [20-22, 27, 28] in this population may influence management strategies, emerging pulsed field ablation may result in increased referral for catheter ablation, given the improved safety profile and efficiency [29].

Sudden Cardiac Death, primarily caused by ventricular arrhythmias, is one of the most common causes of death in COPD patients, with patients often dying at home with no warning signs [3]. It has been suggested that acute COPD exacerbation magnifies airflow obstruction, resulting in stimulation of arterial chemoreceptors and pulmonary stretch receptors, which strengthen sympathetic nerve activity and reduce vagal tone [30]. Konecny et al conducted a retrospective analysis of 2800 patients with COPD determined by pulmonary function testing (PFT) with 24-hour holter monitors [9]. Compared to 13% of patients without COPD, 23% of COPD patients had documented non-sustained VT (p<0.001). The risk of VT increased with COPD severity, and COPD was an independent predictor of all-cause mortality adjusting for left ventricular ejection fraction.

In our study, one patient underwent implantation of an implantable Cardiac defibrillator for sustained ventricular tachycardia, which was documented as a result of continuous monitoring from the ILR. This finding further underscores the importance of considering COPD severity as a nontraditional risk factor when evaluating a patient for primary prevention ICD placement [31]. Nahsuk et al. have previously shown that ICDs reduce mortality in patients with COPD, and COPD is associated with two times the rate of appropriate ICD shocks [5].

Despite the high morbidity and mortality burden conferred by COPD and AF, referred to as the “AF-COPD syndemic,”[26] there exists no effective marker to predict how arrhythmia occurrence influences COPD exacerbation and adverse outcomes. Investigating the potential bidirectional relationship between COPD exacerbations and arrhythmogenesis could provide valuable insights for improving the management of COPD patients at risk for arrhythmias. By increasing awareness about the overlapping symptomatology and potential risk for arrhythmias in COPD patients, we advocate for a multi-disciplinary collaborative approach for referring this underdiagnosed vulnerable population for arrhythmia management.

Our study has several limitations; given the small sample size as part of this pilot study, the power and precision of our effect estimate, given the frequency of the primary outcome, is limited. This is the first analysis using long-term continuous monitoring to evaluate the incidence of arrhythmias in patients with COPD. Therefore, the prevalence of arrhythmias in this population is still being determined. Given the sample’s size, we were unable to correlate acute COPD exacerbation with incident arrhythmias; however, a strength of this analysis is the finding that arrhythmias occur in patients considered stable from a COPD perspective. This was a single-center study, and large-scale prospective clinical trials are needed to validate and generalize the results.

Continuous monitoring was highly effective in diagnosing significant arrhythmic events in patients with moderate-severe COPD. Awareness should be raised about the high arrhythmic risk in this population that is often not referred to electrophysiology, and the role of continuous monitoring should be evaluated in larger studies. The results presented above could expand the indications for ILR implantation in high-risk populations, such as patients with COPD.

## Data Availability

Data available on request from the authors

## Acknowledgments

a) Acknowledgments

None

